# Acute Kidney Injury Among Neonates With Sepsis: Incidence, Predictors and Early Mortality at Hoima Regional Referral Hospital

**DOI:** 10.64898/2025.12.03.25341563

**Authors:** Yasin Ahmed H. Abshir, Bahari Yusuf, Tom Ediamu, Agwu Ezera, Amina Abshir, Theoneste Hakizimana, Abishir Mohamud Hirsi, Abdullahi Abdirizak Farah, Hamdi M. Yusuf, Ahmed Mohamed Nour, Walyeldin Elfakey

**Affiliations:** Department of Pediatrics and Child Health, Kampala International University, Uganda; Faculty of Medicine, University of Bahri, Khartoum Sudan; Department of Internal Medicine, Kampala International University, Uganda; Department of Microbiology and Immunology, Faculty of Biomedical Sciences Kampala International University Western Campus, Ishaka Bushenyi, Uganda; Department of Obstetrics and Gynecology, Kampala International University, Ishaka, Uganda; Faculty of Medicine, International University of Africa, Khartoum Sudan

**Keywords:** Acute kidney injury, Neonatal sepsis, Mortality Predictors, Uganda, KDIGO, Neonate, Hoima Regional Referral Hospital

## Abstract

**Background:** Sepsis is a potentially fatal condition frequently complicated by multi-organ dysfunction, with the kidney being among the most vulnerable organs. Neonatal acute kidney injury (AKI) contributes to prolonged hospitalization and increased mortality. Despite evidence from other countries, data on AKI in neonatal sepsis are scarce in Uganda. This study determined the incidence, Predictors, and early mortality associated with AKI among neonates with sepsis admitted at Hoima Regional Referral Hospital (HRRH).

**Methods:** This was a hospital-based prospective study conducted in the neonatal intensive care unit (NICU) of HRRH. All neonates with clinical or culture-confirmed sepsis whose caregivers consented were enrolled consecutively. AKI was diagnosed using the modified Kidney Disease Improving Global Outcomes (KDIGO) criteria. Serum creatinine levels and urine output were monitored, and participants were followed for 7 days to determine early mortality. Data were analyzed using SPSS version 25. Modified Poisson regression was used to identify independent Predictors, with significance set at *p* < 0.05.

**Results:** A total of 106 neonates were enrolled, of whom 58.5% were female and 82.1% presented within the first 72 hours of life. The incidence of AKI was **20.8% (22/106)**. Independent Predictors for AKI included maternal fever in the week preceding delivery (*p* = 0.004), neonatal convulsions (*p* = 0.011), shock (*p* = 0.002), failure to pass urine in the previous 24 hours (*p* = 0.001), and low birth weight <1.5 kg (*p* = 0.016). The early mortality rate was significantly higher among neonates with AKI (31.8%) compared to those without AKI (2.4%) (*p* < 0.001).

**Conclusion:** AKI is common among neonates with sepsis, occurring in one in every five cases, and is associated with markedly increased mortality. Early identification and management of at-risk neonates—especially those with maternal fever, low birth weight, or shock—are crucial. Strengthening antenatal infection control and neonatal renal monitoring is recommended.

## Background

Sepsis is a potentially fatal clinical condition associated with multiple physiological and biochemical disturbances affecting several vital organs, with the kidney being particularly vulnerable. Acute kidney injury (AKI) in neonates contributes to multi-organ dysfunction syndrome (MODS), prolonged hospital stay, and increased mortality. Globally, an estimated 1.3 million cases of neonatal sepsis occur annually, accounting for about 15% of infant deaths. While sepsis contributes to 9–15% of neonatal deaths in high-income countries, it accounts for up to 27% in resource-limited settings such as sub-Saharan Africa AKI in Neonatal Sepsis.

Neonates are particularly susceptible to AKI because their kidneys are immature and characterized by reduced glomerular filtration rate, increased renal vascular resistance, and poor sodium reabsorption. These factors make them highly vulnerable to hypoperfusion and ischemic injury during systemic infections. In the context of sepsis, inflammatory mediators, altered renal hemodynamics, and exposure to nephrotoxic drugs further exacerbate renal injury.

The Kidney Disease Improving Global Outcomes (KDIGO) criteria define neonatal AKI as an increase in serum creatinine of ≥0.3 mg/dL within 48 hours or 1.5 times baseline within 7 days, or urine output <1 mL/kg/hour for 24 hours.

Studies in various countries have reported neonatal AKI incidence rates ranging from 18% to 74%, depending on population and diagnostic criteria used. For instance, the international AWAKEN study involving over 2,000 neonates reported an incidence of 30%, which was associated with increased mortality and prolonged NICU stay. Other studies have identified sepsis, birth asphyxia, prematurity, and exposure to nephrotoxic drugs as major Predictors for AKI among neonates.

However, data from low-income settings remain limited despite the high burden of neonatal infections and inadequate renal monitoring capacity.

In Uganda, neonatal sepsis remains a major contributor to hospital admissions and mortality, accounting for a significant proportion of the national neonatal death rate of 27 per 1,000 live births. At Hoima Regional Referral Hospital (HRRH), sepsis is one of the most frequent neonatal diagnoses, with 40–50 cases admitted monthly. Yet, renal function assessment is not routinely performed as part of clinical care, and the burden of AKI in this population is not known.

This study therefore aimed to determine the incidence, Predictors, and early mortality associated with acute kidney injury among neonates with sepsis admitted at Hoima Regional Referral Hospital. Understanding these factors will help guide early diagnosis, prevention, and management strategies to improve neonatal outcomes in resource-limited settings.

## Methods

### Study design and setting

A hospital-based prospective observational study was conducted at Hoima Regional Referral Hospital (HRRH), a major referral and teaching facility for Kampala International University in mid-western Uganda. The study took place in the Neonatal Intensive Care Unit (NICU), which manages inborn and referred neonates with conditions such as sepsis, asphyxia, and prematurity. Data were collected from March to June 2025.

### Study population and eligibility

All neonates aged 0–28 days admitted with clinical or laboratory-confirmed sepsis were eligible. Sepsis was defined by typical clinical features and abnormal total leukocyte counts (<5,000/mm³ or >30,000/mm³). Neonates with congenital renal anomalies or those who died before sampling were excluded. Written informed consent was obtained from parents or guardians.

### Sample size and sampling

For objective one, the sample size was calculated using Daniel’s formula: 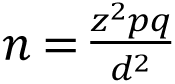 Using findings by Palle et al. (2020) who reported that the incidence of acute kidney injury among neonates with sepsis was 6.5%, we set *p = 0.065*, *q = 1 – p = 0.935*, *d = 0.05*, and *Z = 1.96* for a 95% confidence level. Substituting these values, the calculated sample size was n = 93.

For Objective two, Epi-info online sample size calculator was used (https://www.openepi.com/SampleSize/SSCohort.htm). Using findings by (Gabri et al., 2021) who reported that AKI was found in 47.8% of neonates with sepsis confirmed by culture and sensitivity as opposed to the 77.2% among those in whom it was not confirmed by culture, for a power of 80% and 95% level of confidence, the sample size required was 96 using Fleiss method.

For objective three, Epi-info online sample size calculator was used (https://www.openepi.com/SampleSize/SSCohort.htm). Using findings by (Pradhan et al., 2014) who reported that compared to neonates without AKI (24.1%), septic newborns with AKI had a greater mortality rate (54.5%) (Pradhan et al., 2014), for a power of 80% and 95% level of confidence, the sample size required was 92 using Fleiss method.

Taking the bigger sample size (96) and adding 10% to cater for loss of follow up, the sample size required was 106.

### Data collection and study procedure

Data were obtained using a structured, pretested questionnaire capturing socio-demographic, perinatal, and clinical information. The principal investigator and trained nurses collected data through caregiver interviews, record review, and physical examination. Each neonate was stabilized according to HRRH protocols, then had 5 mL of venous blood drawn for complete blood count and serum creatinine (SCr) measurement. Baseline SCr was taken within 24 hours of admission and repeated on day 3 and day 7. Urine output was assessed based on caregiver-reported history.

Acute kidney injury was defined using modified KDIGO criteria—an increase in SCr ≥ 0.3 mg/dL within 48 h, or ≥ 1.5 × baseline within 7 days.

All neonates received standard sepsis management with empiric antibiotics, fluids, and supportive care. Each neonate was followed for seven days to determine Mortality.

### Laboratory procedure

Serum creatinine was measured using the Jaffe’s kinetic colorimetric method on a Mindray BS-240 analyzer. Venous blood was centrifuged at 3,000 rpm for 10 minutes to obtain serum. Internal quality control using normal and high creatinine standards was performed daily. Sepsis was further supported by hematological results and, where possible, blood cultures.

### Data management and analysis

Data were entered in EpiData 3.1 and analyzed using SPSS 25. Descriptive statistics summarized baseline characteristics. Bivariate analysis identified potential predictors of AKI; variables with *p* < 0.2 were included in a multivariable modified Poisson regression model. Statistical significance was set at *p* < 0.05, and results were reported as adjusted relative risks (aRR) with 95% confidence intervals. Mortality outcomes were assessed using the Chi-square test to determine associations with AKI. Statistical significance was set at p < 0.05, and results were reported as adjusted relative risks (aRR) with 95% confidence intervals.

### Ethical considerations

Ethical approval was obtained from the Kampala International University Research and Ethics Committee (KIU-REC Ref: KIU-2025-767) and permission from the HRRH Medical Director. Written informed consent was obtained from caregivers, and confidentiality maintained using coded identifiers. All procedures followed the Declaration of Helsinki.

## Results

### Baseline characteristics of the study participants

In this study, we enrolled 106 neonates with neonatal sepsis. Slightly over half (58.5%) were female. Majority (82.1%) presented with in the first 72 hours and therefore considered to have had early onset sepsis. Only 40.6% had been born at term according to their gestational age. Close to one third (27.4%) had an Apgar score less than 7 at the 5^th^ minute. The details so the baseline characteristics are shown in table 1 below.

**Table 1:** Baseline characteristics of study participants.

| Characteristic | Frequency | Percentage |
| --- | --- | --- |
| **Sex** |  |  |
| Male | 44 | 41.5 |
| Female | 62 | 58.5 |
| **Age** |  |  |
| ≤72 hrs. | 87 | 82.1 |
| 73+ hrs. | 19 | 17.9 |
| **UTI in pregnancy** |  |  |
| No | 52 | 49.1 |
| Yes | 54 | 50.9 |
| **Maternal fevers** |  |  |
| No | 66 | 62.3 |
| Yes | 40 | 37.7 |
| **Difficulty breathing** |  |  |
| No | 36 | 34.0 |
| Yes | 70 | 66.0 |
| **Altered consciousness** |  |  |
| No | 84 | 79.2 |
| Yes | 22 | 20.8 |
| **Dehydrated** |  |  |
| No | 98 | 92.5 |
| Yes | 8 | 7.5 |
| **Convulsed** |  |  |
| No | 83 | 78.3 |
| Yes | 23 | 21.7 |
| **Vomiting** |  |  |
| No | 102 | 96.2 |
| Yes | 4 | 3.8 |
| **Jaundice** |  |  |
| No | 74 | 69.8 |
| Yes | 32 | 30.2 |
| **Tachycardia** |  |  |
| No | 26 | 24.5 |
| Yes | 80 | 75.5 |
| **Shock** |  |  |
| No | 91 | 85.8 |
| Yes | 15 | 14.2 |
| **Membrane rupture duration** |  |  |
| ≤18hrs | 34 | 32.1 |
| 18+ hrs. | 72 | 67.9 |
| **Gestational age** |  |  |
| ≤32 | 29 | 27.4 |
| 33-36 | 34 | 32.1 |
| 37+ | 43 | 40.6 |
| **Passed urine in last 24 hrs.** |  |  |
| No | 54 | 50.9 |
| Yes | 52 | 49.1 |
| **Birth weight (kg)** |  |  |
| <1.5 | 17 | 16.0 |
| 1.5-2.4 | 55 | 51.9 |
| 2.5+ | 34 | 32.1 |
| **APGAR score** |  |  |
| < 7.0 | 29 | 27.4 |
| 7.0+ | 77 | 72.6 |
| **APH** |  |  |
| No | 101 | 95.3 |
| Yes | 5 | 4.7 |
| **Invasive procedure** |  |  |
| No | 93 | 87.7 |
| Yes | 13 | 12.3 |
| **Hemoglobin** |  |  |
| Normal | 47 | 44.3 |
| Low | 50 | 47.2 |
| High | 9 | 8.5 |
| **Platelet count** |  |  |
| Normal | 40 | 37.7 |
| Low | 31 | 29.2 |
| High | 35 | 33.0 |
| **Leukocytes** |  |  |
| Normal | 40 | 37.7 |
| Low | 32 | 30.2 |
| High | 34 | 32.1 |

*UTI=Urinary tract infection, APH=Antepartum hemorrhage.*

## Incidence of acute kidney injury

### 4.2 Incidence of acute kidney injury among neonates with sepsis admitted at HRRH

Of the 106 neonates enrolled with neonatal sepsis, 22 had acute kidney injury, representing an incidence of 20.8% with corresponding 95% confidence interval of 13.2-29.2% as shown in figure 1 below.

**Figure 1:**
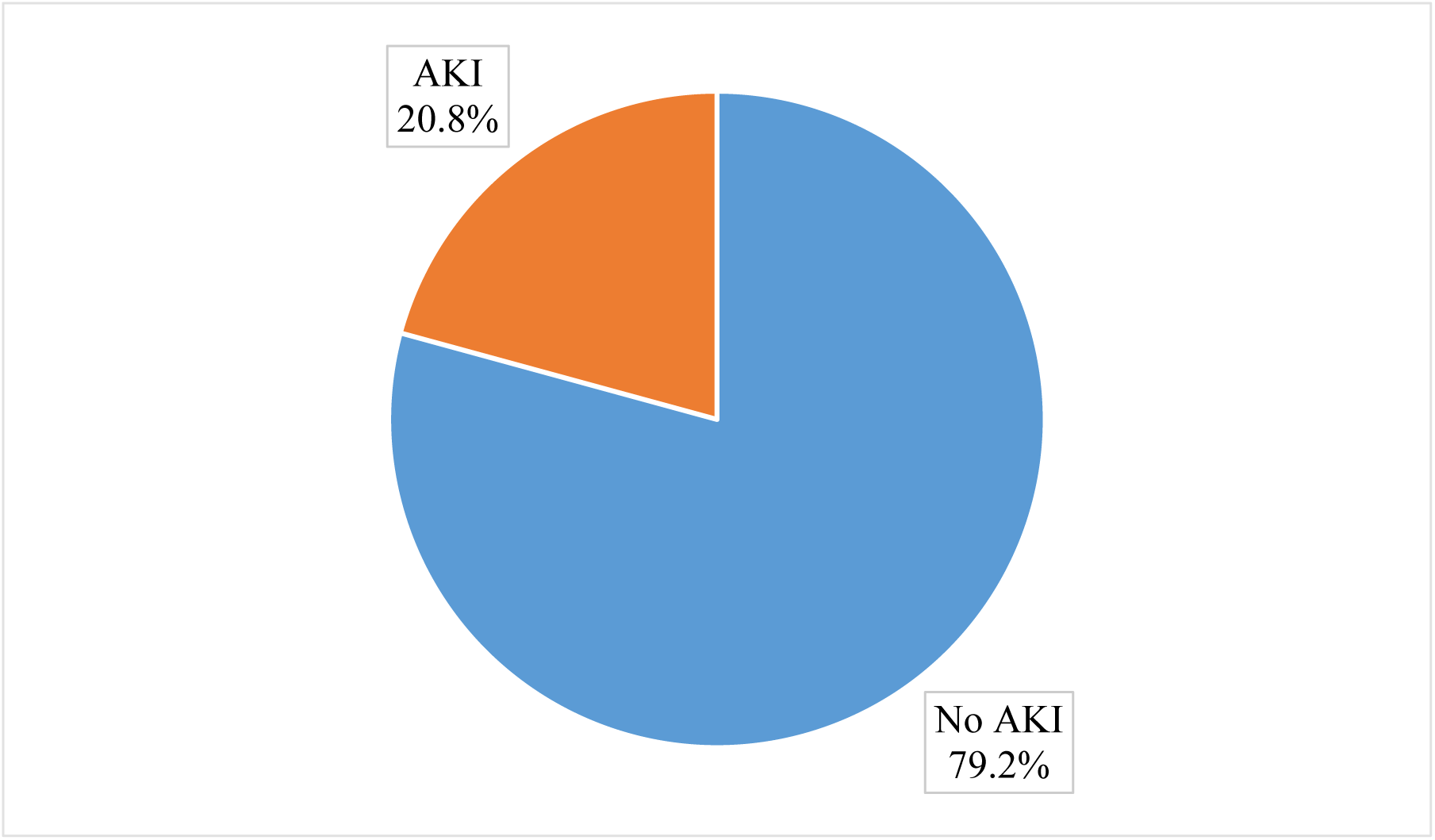
Incidence of acute kidney injury among neonates with sepsis admitted at HRRH.

### Risk factors of acute kidney injury among neonates with sepsis admitted at HRRH

The variables considered for multivariate analysis (P<0.2) were sex, maternal fever, altered level of consciousness, dehydration, convulsions, shock, gestational age, not passing urine in the preceding 24 hours, birth weight and Apgar score. The details of the bivariate analysis are shown in table 2 below.

**Table 2:**
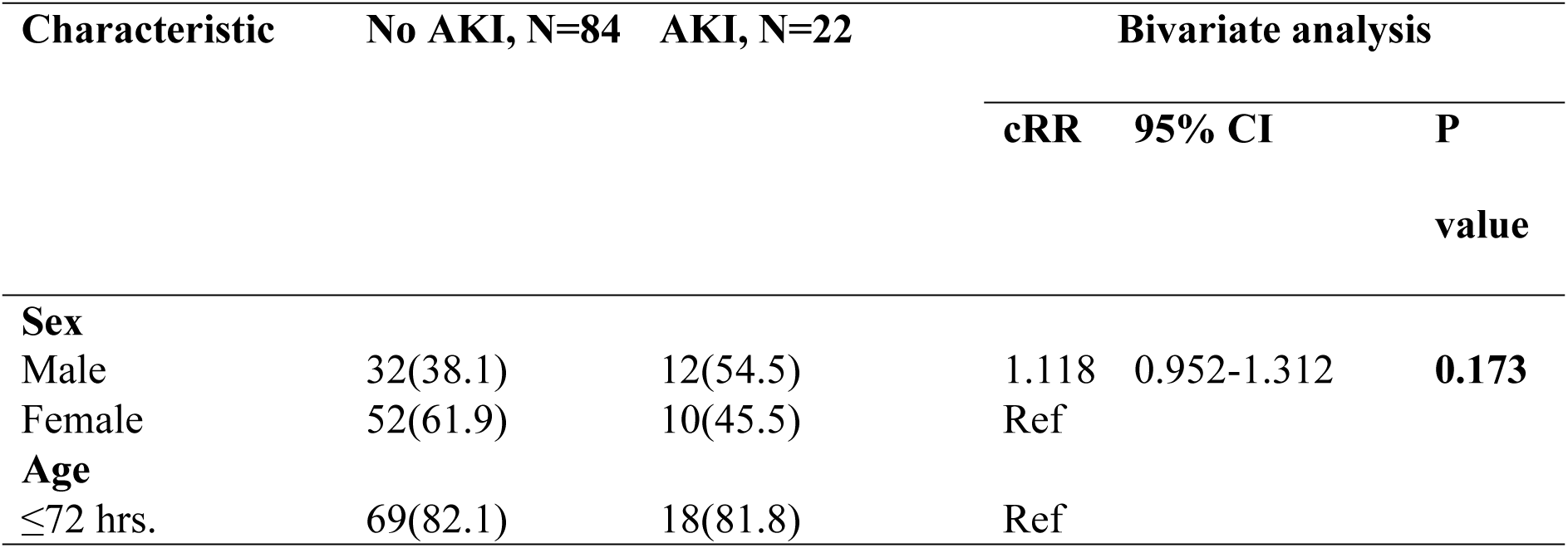
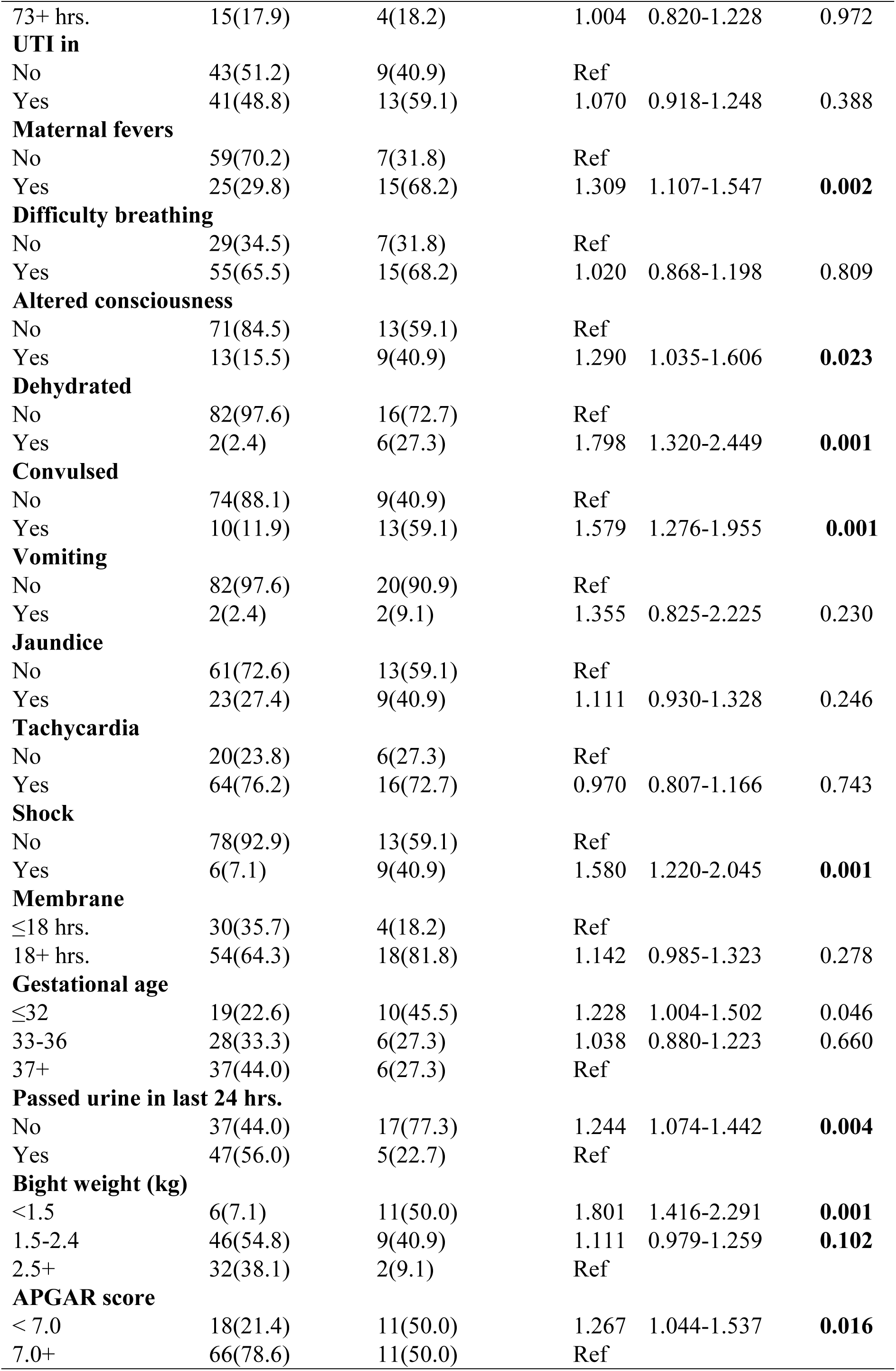
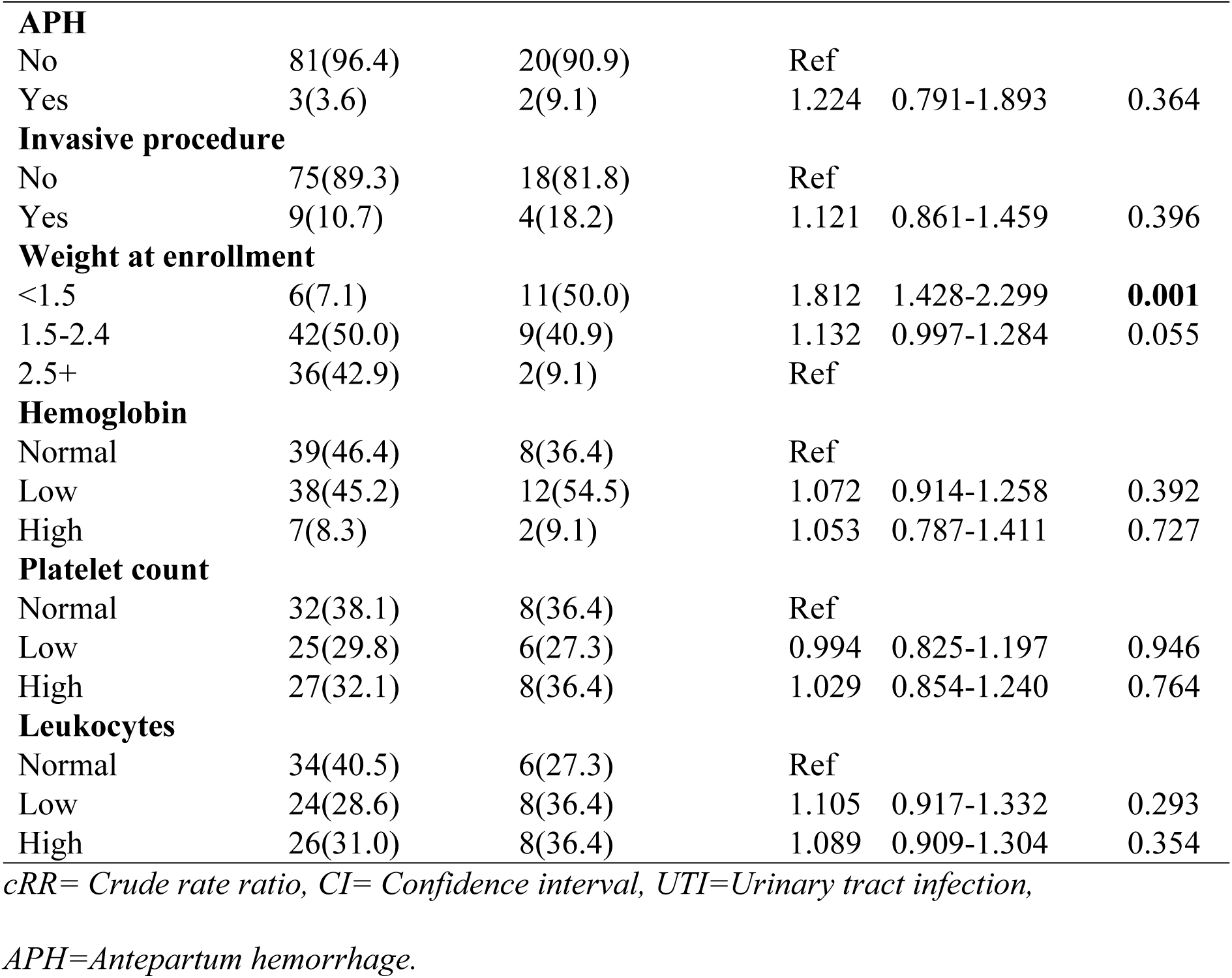
Bivariate analysis of risk factors of acute kidney injury among neonates with sepsis admitted at HRRH.

| Characteristic | No AKI, N=84 | AKI, N=22 | Bivariate analysis |  |  |
| --- | --- | --- | --- | --- | --- |
|  |  |  | cRR | 95% CI | P value |
| **Sex** |  |  |  |  |  |
| Male | 32(38.1) | 12(54.5) | 1.118 | 0.952-1.312 | **0.173** |
| Female | 52(61.9) | 10(45.5) | Ref |  |  |
| **Age** |  |  |  |  |  |
| ≤72 hrs. | 69(82.1) | 18(81.8) | Ref |  |  |
| 73+ hrs. | 15(17.9) | 4(18.2) | 1.004 | 0.820-1.228 | 0.972 |
| **UTI in** |  |  |  |  |  |
| No | 43(51.2) | 9(40.9) | Ref |  |  |
| Yes | 41(48.8) | 13(59.1) | 1.070 | 0.918-1.248 | 0.388 |
| **Maternal fevers** |  |  |  |  |  |
| No | 59(70.2) | 7(31.8) | Ref |  |  |
| Yes | 25(29.8) | 15(68.2) | 1.309 | 1.107-1.547 | **0.002** |
| **Difficulty breathing** |  |  |  |  |  |
| No | 29(34.5) | 7(31.8) | Ref |  |  |
| Yes | 55(65.5) | 15(68.2) | 1.020 | 0.868-1.198 | 0.809 |
| **Altered consciousness** |  |  |  |  |  |
| No | 71(84.5) | 13(59.1) | Ref |  |  |
| Yes | 13(15.5) | 9(40.9) | 1.290 | 1.035-1.606 | **0.023** |
| **Dehydrated** |  |  |  |  |  |
| No | 82(97.6) | 16(72.7) | Ref |  |  |
| Yes | 2(2.4) | 6(27.3) | 1.798 | 1.320-2.449 | **0.001** |
| **Convulsed** |  |  |  |  |  |
| No | 74(88.1) | 9(40.9) | Ref |  |  |
| Yes | 10(11.9) | 13(59.1) | 1.579 | 1.276-1.955 | **0.001** |
| **Vomiting** |  |  |  |  |  |
| No | 82(97.6) | 20(90.9) | Ref |  |  |
| Yes | 2(2.4) | 2(9.1) | 1.355 | 0.825-2.225 | 0.230 |
| **Jaundice** |  |  |  |  |  |
| No | 61(72.6) | 13(59.1) | Ref |  |  |
| Yes | 23(27.4) | 9(40.9) | 1.111 | 0.930-1.328 | 0.246 |
| **Tachycardia** |  |  |  |  |  |
| No | 20(23.8) | 6(27.3) | Ref |  |  |
| Yes | 64(76.2) | 16(72.7) | 0.970 | 0.807-1.166 | 0.743 |
| **Shock** |  |  |  |  |  |
| No | 78(92.9) | 13(59.1) | Ref |  |  |
| Yes | 6(7.1) | 9(40.9) | 1.580 | 1.220-2.045 | **0.001** |
| **Membrane** |  |  |  |  |  |
| ≤18 hrs. | 30(35.7) | 4(18.2) | Ref |  |  |
| 18+ hrs. | 54(64.3) | 18(81.8) | 1.142 | 0.985-1.323 | 0.278 |
| **Gestational age** |  |  |  |  |  |
| ≤32 | 19(22.6) | 10(45.5) | 1.228 | 1.004-1.502 | 0.046 |
| 33-36 | 28(33.3) | 6(27.3) | 1.038 | 0.880-1.223 | 0.660 |
| 37+ | 37(44.0) | 6(27.3) | Ref |  |  |
| **Passed urine in last 24 hrs.** |  |  |  |  |  |
| No | 37(44.0) | 17(77.3) | 1.244 | 1.074-1.442 | **0.004** |
| Yes | 47(56.0) | 5(22.7) | Ref |  |  |
| **Bight weight (kg)** |  |  |  |  |  |
| <1.5 | 6(7.1) | 11(50.0) | 1.801 | 1.416-2.291 | **0.001** |
| 1.5-2.4 | 46(54.8) | 9(40.9) | 1.111 | 0.979-1.259 | **0.102** |
| 2.5+ | 32(38.1) | 2(9.1) | Ref |  |  |
| **APGAR score** |  |  |  |  |  |
| < 7.0 | 18(21.4) | 11(50.0) | 1.267 | 1.044-1.537 | **0.016** |
| 7.0+ | 66(78.6) | 11(50.0) | Ref |  |  |
| **APH** |  |  |  |  |  |
| No | 81(96.4) | 20(90.9) | Ref |  |  |
| Yes | 3(3.6) | 2(9.1) | 1.224 | 0.791-1.893 | 0.364 |
| **Invasive procedure** |  |  |  |  |  |
| No | 75(89.3) | 18(81.8) | Ref |  |  |
| Yes | 9(10.7) | 4(18.2) | 1.121 | 0.861-1.459 | 0.396 |
| **Weight at enrollment** |  |  |  |  |  |
| <1.5 | 6(7.1) | 11(50.0) | 1.812 | 1.428-2.299 | **0.001** |
| 1.5-2.4 | 42(50.0) | 9(40.9) | 1.132 | 0.997-1.284 | 0.055 |
| 2.5+ | 36(42.9) | 2(9.1) | Ref |  |  |
| **Hemoglobin** |  |  |  |  |  |
| Normal | 39(46.4) | 8(36.4) | Ref |  |  |
| Low | 38(45.2) | 12(54.5) | 1.072 | 0.914-1.258 | 0.392 |
| High | 7(8.3) | 2(9.1) | 1.053 | 0.787-1.411 | 0.727 |
| **Platelet count** |  |  |  |  |  |
| Normal | 32(38.1) | 8(36.4) | Ref |  |  |
| Low | 25(29.8) | 6(27.3) | 0.994 | 0.825-1.197 | 0.946 |
| High | 27(32.1) | 8(36.4) | 1.029 | 0.854-1.240 | 0.764 |
| **Leukocytes** |  |  |  |  |  |
| Normal | 34(40.5) | 6(27.3) | Ref |  |  |
| Low | 24(28.6) | 8(36.4) | 1.105 | 0.917-1.332 | 0.293 |
| High | 26(31.0) | 8(36.4) | 1.089 | 0.909-1.304 | 0.354 |

*cRR= Crude rate ratio, CI= Confidence interval, UTI=Urinary tract infection,*
*APH=Antepartum hemorrhage.*

In the multivariable analysis, the independent risk factors of acute kidney injury among neonates with sepsis were: history of maternal fevers (aRR=1.161, CI=1.001-1.347, P=0.048), history of convulsions (aRR=1.364, CI=1.126-1.651, P=0.001), presence of shock (aRR=1.171, CI=1.057-1.598, P=0.022), not passing urine in the preceding 24 hours (aRR=1.193, CI=1.053-1.351, P=0.005) and birth weight less than 1.5 kg (aRR=1.440, CI=1.119-1.853, P=0.005). The rate of acute kidney injury was increased by 16.1% among neonates whose mothers had a history of fevers before delivery, by 36.4% among neonates with a history of convulsions, by 17.1% among neonates who were found to be in shock on examination, by 19.3% among those who had not passed urine in the preceding 24 hours and by 44.0% among those whose birth weight was less than 1.5 kg. The details of the multivariate analysis are shown in table 3 below.

**Table 3:** Multivariable analysis of risk factors of acute kidney injury among neonates with sepsis admitted at HRRH.

| Characteristic | Bivariate analysis |  |  | Multivariate analysis |  |  |
| --- | --- | --- | --- | --- | --- | --- |
|  | cRR | 95% CI | P value | aRR | 95% CI | P value |
| **Sex** |  |  |  |  |  |  |
| Male | 1.118 | 0.952-1.312 | 0.173 | 1.055 | 0.943-1.180 | 0.351 |
| Female | Ref |  |  |  |  |  |
| **Maternal fevers** |  |  |  |  |  |  |
| No | Ref |  |  |  |  |  |
| Yes | **1.309** | **1.107-1.547** | **0.002** | **1.161** | **1.001-1.347** | **0.048** |
| **Altered consciousness** |  |  |  |  |  |  |
| No | Ref |  |  |  |  |  |
| Yes | 1.290 | 1.035-1.606 | 0.023 | 1.133 | 0.954-1.345 | 0.154 |
| **Dehydrated** |  |  |  |  |  |  |
| No | Ref |  |  |  |  |  |
| Yes | 1.798 | 1.320-2.449 | <0.001 | 1.298 | 0.320-2.449 | 0.061 |
| **Convulsed** |  |  |  |  |  |  |
| No | Ref |  |  |  |  |  |
| Yes | **1.579** | **1.276-1.955** | **<0.001** | **1.364** | **1.126-1.651** | **0.001** |
| **Shock** |  |  |  |  |  |  |
| No | Ref |  |  |  |  |  |
| Yes | **1.580** | **1.220-2.045** | **0.001** | **1.171** | **1.057-1.598** | **0.022** |
| **Gestational age** |  |  |  |  |  |  |
| ≤32 | 1.228 | 1.004-1.502 | 0.046 | 1.067 | 0.865-1.317 | 0.545 |
| 33-36 | 1.038 | 0.880-1.223 | 0.660 | 1.037 | 0.890-1.209 | 0.642 |
| 37+ | Ref |  |  |  |  |  |
| **Passed urine in last 24 hrs** |  |  |  |  |  |  |
| No | **1.244** | **1.074-1.442** | **0.004** | **1.193** | **1.053-1.351** | **0.005** |
| Yes | Ref |  |  |  |  |  |
| **Bight weight (kg)** |  |  |  |  |  |  |
| <1.5 | **1.801** | **1.416-2.291** | **<0.001** | **1.440** | **1.119-1.853** | **0.005** |
| 1.5-2.4 | 1.111 | 0.979-1.259 | 0.102 | 1.062 | 0.929-1.215 | 0.379 |
| 2.5+ | Ref |  |  |  |  |  |
| **APGAR score** |  |  |  |  |  |  |
| < 7.0 | 1.267 | 1.044-1.537 | 0.016 | 1.172 | 0.969-1.416 | 0.102 |
| 7.0+ | Ref |  |  |  |  |  |

*cRR= Crude rate ratio, aRR=adjusted rate ratio, CI= Confidence interval.*

### Early mortality rate among the neonates with AKI and sepsis in comparison with those without AKI admitted at HRRH

Of the 106 study participants, 9 passed away representing an overall mortality rate of 8.5%, with a 95% confidence interval of 2.8-15.1%. The mortality was significantly higher in the neonates that had AKI, with a mortality of 31.8% versus 2.4% among those who did not have AKI, and a chi squire p value of <0.001. Of these deaths, 33% occurred on the second day of life, while 66% occurred on the 5^th^ of life. The details on mortality are shown in table 4 below.

**Table 4:** Early Mortality Outcome among Neonates with Sepsis by AKI Status (N = 106)

| **Outcome** | **AKI**<br>**(N=22)** | **No AKI,**<br>**(N=84)** | **Overall**<br>**(N=106)** | **P value** |
| --- | --- | --- | --- | --- |
| **Mortality** |  |  |  | **<0.001** |
| Died | 7(31.8%) | 2(2.4%) | 9(8.5%) |  |
| Survived | 15(68.2%) | 82(97.6%) | 97(91.5%) |  |

## Discussion

### Overview

This study determined the incidence, Predictors, and early mortality associated with acute kidney injury (AKI) among neonates with sepsis admitted to Hoima Regional Referral Hospital. The findings show that AKI occurred in 20.8% of septic neonates, and that affected neonates had a markedly higher mortality rate (31.8%) compared to those without AKI (2.4%). The independent predictors of AKI were maternal fever before delivery, neonatal convulsions, presence of shock, anuria within 24 hours, and birth weight below 1.5 kg. These findings highlight that AKI is a significant complication of neonatal sepsis in Uganda, contributing substantially to poor early outcomes.

### Incidence of acute kidney injury

The incidence of AKI in this study (20.8%) is within the range reported internationally, where AKI prevalence among septic neonates has varied from 18% to 36% depending on population and diagnostic criteria. The proportion observed in this study is comparable to that reported in the United States (20%) by Grundmeier et al. (2020) and slightly lower than the 36.1% reported in Kenya by Munyendo et al. (2023). This variation may be attributed to differences in diagnostic methods, timing of serum creatinine measurement, and severity of sepsis. Studies in India also reported incidences of 24% (Durga & Rudrappa, 2017) and 27.5% (Pradhan et al., 2014), consistent with our findings. The present study’s relatively lower incidence compared to Kenya may reflect early diagnosis, improved infection control, or differences in neonatal admission criteria at Hoima Regional Referral Hospital.

The observed one-in-five burden underscores that AKI remains a common complication among neonates with sepsis, even in settings where renal monitoring is not routinely performed. This finding provides local evidence supporting the inclusion of renal function assessment in the routine evaluation of septic neonates.

### Predictors for acute kidney injury

Several maternal and neonatal factors were associated with AKI in this study. Maternal fever before delivery emerged as an independent predictor, consistent with findings from Kenya (Munyendo et al., 2023) and India (Durga & Rudrappa, 2017), which reported that maternal infections and prolonged rupture of membranes predispose neonates to sepsis and subsequent AKI. Fever during pregnancy likely reflects intrauterine infection that compromises renal perfusion and increases inflammatory cytokines in the neonate.

Neonatal convulsions were also independently associated with AKI. This relationship is likely due to hypoxic-ischemic injury during convulsive episodes, which reduces renal blood flow and precipitates acute tubular necrosis. This finding aligns with reports by Nickavar et al. (2017) and Pradhan et al. (2014), who observed higher AKI rates among neonates with neurological complications and disseminated intravascular coagulation.

Shock was another strong predictor, similar to studies from India and Iran (Pradhan et al., 2014; Nickavar et al., 2017), where septic shock was identified as a key driver of renal hypoperfusion and ischemia. The pathophysiological mechanism involves reduced renal perfusion pressure and systemic inflammatory injury to renal microvasculature.

Additionally, failure to pass urine within 24 hours was a clear marker of AKI, as oligo-anuria is one of the earliest clinical indicators of renal dysfunction. This aligns with the KDIGO criteria used globally for neonatal AKI diagnosis. Low birth weight (<1.5 kg) was also independently associated with AKI, consistent with evidence that preterm and small neonates have immature kidneys with reduced nephron endowment, making them highly susceptible to injury (Coleman et al., 2022).

These associations emphasize that both maternal infections and neonatal hemodynamic instability play critical roles in the development of AKI. Early recognition of these Predictors may allow for timely interventions to prevent renal injury and improve outcomes.

### Early mortality and outcomes

The early mortality rate among neonates with sepsis was 7.5%, but was dramatically higher in those with AKI (31.8%) compared to those without (2.4%), a statistically significant difference (p < 0.001). This confirms that AKI is a major predictor of early death among septic neonates. Similar findings were observed in studies from Iran (15.4% vs 4%) (Nickavar et al., 2017) and the United States, where AKI tripled the risk of 30-day mortality (Grundmeier et al., 2020). The higher mortality in this study may reflect delays in presentation, limited renal replacement therapy, and resource constraints in monitoring critically ill neonates.

These results reinforce that AKI not only complicates the course of sepsis but also significantly worsens short-term outcomes. The presence of AKI likely reflects severe systemic illness and multi-organ dysfunction. Early recognition and aggressive management of shock and dehydration, along with avoidance of nephrotoxic agents, are therefore crucial to reducing mortality.

The findings of this study are consistent with the broader literature showing that neonatal AKI is a frequent and severe complication of sepsis in both high- and low-income settings. The incidence observed aligns closely with multicenter studies such as the AWAKEN (Assessment of the worldwide AKI epidemiology in newborns)cohort, which reported an AKI incidence of 30% and linked it to prolonged hospital stay and mortality. However, this study provides the first local evidence from a Ugandan referral hospital quantifying the burden of AKI among septic neonates and identifying modifiable predictors.

The strong association between AKI and early mortality highlights the need to integrate renal monitoring (serum creatinine and urine output tracking) into neonatal sepsis protocols at regional hospitals. Routine maternal infection screening during antenatal care and prompt management of neonatal shock could further prevent AKI-related deaths.

### Strengths and limitations

This study is among the few prospective studies on neonatal AKI in Uganda and the first to evaluate the incidence and outcomes of AKI among neonates with sepsis in Hoima. Its prospective design and use of standardized KDIGO criteria enhance the reliability of results. However, the study was limited by its single-center design and relatively small sample size, which may affect generalizability. Additionally, renal ultrasound and urine chemistry were not performed due to limited resources.

## Conclusion

This study found that acute kidney injury (AKI) occurred in 20.8% of neonates with sepsis admitted to Hoima Regional Referral Hospital. AKI was strongly associated with a significantly higher risk of early mortality, with deaths occurring in nearly one-third of affected neonates. The independent predictors of AKI included maternal fever before delivery, neonatal convulsions, presence of shock, failure to pass urine within 24 hours, and low birth weight below 1.5 kg. These findings demonstrate that AKI is a common and serious complication of neonatal sepsis and an important contributor to early neonatal deaths in resource-limited settings.

Early identification of high-risk neonates, close monitoring of renal function, and prompt management of sepsis and shock can substantially reduce AKI incidence and mortality. Integration of renal function testing (serum creatinine and urine output monitoring) into neonatal care protocols is crucial for early diagnosis and intervention.

### Recommendations

Routine screening for AKI should be incorporated into the management of all neonates admitted with sepsis, particularly those with maternal fever, low birth weight, or hemodynamic instability. Healthcare providers should strengthen renal monitoring through regular serum creatinine testing and strict urine output charting.

Training neonatal staff on the early recognition of AKI and avoidance of nephrotoxic drugs should be prioritized. At the community level, efforts should focus on preventing maternal infections during pregnancy through antenatal education and infection control measures. Further multicenter studies with larger sample sizes are recommended to explore the long-term renal outcomes of neonates who survive sepsis-associated AKI in Uganda.

**List of abbreviations**
AKI: Acute Kidney Injury,
KDIGO: Kidney Disease: Improving Global Outcomes
SPSS: Statistical Package for the Social Sciences
HRRH: Hoima Regional Referral Hospital

## Ethical considerations and consent

All methods were carried out in accordance with relevant guidelines and regulations. Ethical approval was obtained from the Research and Ethics Committee of Kampala International University Western Campus (Ref No: KIU-2024-767). Written informed consent was obtained from all parents or guardians of the study participants.

## Consent for publication

Not applicable

## Availability of data and materials

Data is available upon request. Requests should be sent to (YAHA)

## Author contribution

**YAHA** was the principle investigator, conceived and designed the study, collected data, analysed data and wrote the draft of the manuscript. **WE, TE, EA, TH and AMH** supervised the work and revised the manuscript, **AA**, **AAF, BY, AMA, AMN and HMY** participated in data collection, revised the manuscript and all authors approved the final paper.

## Data Availability

Availability of data and materials: Data is available upon request. Requests should be sent to (YAHA)

## Acknowledgements

Authors thank research assistants and all the participants of the study.

The authors express sincere gratitude to the management and staff of Hoima Regional Referral Hospital, the neonatal unit nurses, and all caregivers who participated in the study. Special appreciation is extended to the Department of Pediatrics and Child Health, Kampala International University, for continuous academic and logistical support.

